# Integrated SARS-CoV-2 serological and virological screening across an acute fever surveillance platform to monitor temporal changes in anti-spike antibody levels and risk of infection during sequential waves of variant transmission — Dominican Republic, March 2021 to August 2022

**DOI:** 10.1101/2022.10.24.22281399

**Authors:** Eric J. Nilles, Michael de St. Aubin, Devan Dumas, William Duke, Marie Caroline Etienne, Gabriela Abdalla, Petr Jarolim, Timothy Oasan, Salome Garnier, Naomi Iihoshi, Beatriz Lopez, Lucia de la Cruz, Yosanly Cornelio Puello, Margaret Baldwin, Kathryn W. Roberts, Farah Peña, Kara Durski, Isaac Miguel Sanchez, Sarah M. Gunter, Alexander R. Kneubehl, Kristy O. Murray, Allison Lino, Sarah Strobel, Amado Alejandro Baez, Colleen L. Lau, Adam Kucharski, Emily Zielinski Gutiérrez, Ronald Skewes-Ramm, Marietta Vasquez, Cecilia Then Paulino

## Abstract

The global SARS-CoV-2 immune landscape and population protection against emerging variants is largely unknown. We assessed SARS-CoV-2 antibody changes in the Dominican Republic and implications for immunological protection against variants of concern. Between March 2021 and August 2022, 2,300 patients with undifferentiated febrile illnesses were prospectively enrolled. Sera was tested for total anti-spike antibodies and simultaneously collected nasopharyngeal samples for acute SARS-CoV-2 infection with RT-PCR. Geometric mean anti-spike titers increased from 6.6 BAU/ml (95% CI 5.1–8.7) to 1,332 BAU/ml (1055–1,682). Multivariable binomial odds ratios for acute SARS-CoV-2 infection were 0.55 (0.40–0.74), 0.38 (0.27–0.55), and 0.27 (0.18–0.40) for the second, third, and fourth versus the first anti-S quartile, with similar findings by viral strain. Integrated serological and virological screening can leverage existing acute fever surveillance platforms to monitor population-level immunological markers and concurrently characterize implications for emergent variant transmission in near real-time.

## Background

Given widespread unreported SARS-CoV-2 infections, variable immunological response based on host immunogenicity, vaccine type, viral strain, timing and sequence of vaccine or viral exposure, and humoral waning, the global SARS-CoV-2 immune landscape is largely unknown. Most countries launched national COVID-19 vaccination campaigns during early 2021, but few studies have characterized population-level immunological responses to SARS-CoV-2 and fewer have aimed to translate findings to immunological protection. Many large national seroepidemiological studies were conducted in the pre-COVID-19 vaccine era and prior to emerging variants of concern, focusing primarily on seroprevalence, i.e., the presence or absence of anti-SARS-CoV-2 antibodies, but not the quantification of antibody levels.^1–4^ This was largely due to an urgent need to understand population-level transmission and transmission risks, but it was also due to a limited understanding of what binding antibody levels mean for immunological protection, and if quantification of binding antibodies translate into actionable or otherwise useful data. While neutralizing antibodies are the generally accepted standard correlate of protection against symptomatic infection,^5–7^ measurement of neutralizing activity is slow and resource intensive and therefore impractical for most population-based studies, particularly in low and middle income countries. Recent approaches have combined screening subsets of populations for neutralizing activity and applying machine learning methods to estimate population-level immunological protection,^8^ but these approaches still require neutralization testing of a certain fraction of samples in addition to the application of machine learning methods. The direct use of binding antibodies to estimate immunological protection is, therefore, attractive, at least for population-based studies where the tolerance for imprecision may be higher than vaccine efficacy trials. While global health authorities including the World Health Organization (WHO) previously cautioned against the use of binding antibodies to assess immunological protection, several large studies subsequently demonstrated that SARS-CoV-2 anti-spike binding antibodies (anti-S) largely track with protection against infection.^5–7^ However, these studies were conducted in the setting of controlled vaccine efficacy studies and prior to the emergence of highly immune evasive viral variants so the utility of anti-S antibodies for understanding immunological protection in a real world setting when transmission is driven by Omicron derived strains is unknown.

Given these knowledge gaps, which we believe are essential for informing and prioritizing public health activities moving forward, we conducted a proof-of-concept study utilizing a novel methodological approach to first characterize population-level temporal changes in anti-S binding antibody titers and second evaluate the utility of anti-S binding antibodies for assessing risk of acute SARS-CoV-2 infection across viral variants and strains.

## Methods

### Setting

The Dominican Republic is an upper middle income Latin American country that shares the island of Hispaniola with Haiti. With almost 11 million residents, it is the second most populous country in the Caribbean.^9,10^ The first laboratory confirmed case of SARS-CoV-2 was reported in the Dominican Republic on 1 March 2020, and strict public health measures commensurate with most regional countries were implemented.^11^ Six discrete waves of SARS-CoV-2 transmission have been observed between March 2020 and August 2022 with the last three predominantly due to B.1.617.2 (Delta, October through November 2021), BA.1 (Omicron, January through February 2022) and post-BA.1 Omicron variants including BA.2, BA.4, and BA.5 (June through August 2022). Peak national cases reported per day were 4-5 times higher during the BA.1 wave (around 6,000 case per day) than during the other waves (around 1,100 to 1,300 cases per day).^6^ A national COVID-19 vaccination campaign was launched in late February 2021 and by 22 March 2021, the start of the current study, approximately 7.4% of the national population had received one COVID-19 vaccine dose.^12^ The principal COVID-19 vaccines administered included the inactivated viral CoronaVac (Sinovac), the adenovirus vector ChAdOx1-S (Oxford/AstraZeneca) and mRNA BNT162b2 (Pfizer/BioNTech) vaccines. Latin America emerged as a global SARS-CoV-2 hotspot early in the COVID-19 pandemic, with model estimates suggesting that by November 2021 the regional cumulative population infected was 57·4% (51·7–63·1).^13^ A national cross-sectional household serological survey in the Dominican Republic estimated that by August 2021, 85.0% (CI 82.1–88.0) of the ≥ 5-year-old population had been immunologically exposed through vaccination, infection or both, and 77.5% (CI 71.3–83) had been previously infected.^8^

### Study design, study sites, participant selection, and ethical considerations

Prospective enrollment was conducted across two study sites: Hospital Dr. Antonio Musa, located in San Pedro de Macoris province in the southeast of the country, and Dr. Toribio Bencosme Hospital in Espaillat province in the northwest of the country. These study sites are part of a longitudinal US CDC-funded acute febrile infection enhanced surveillance platform, which, in collaboration with the Ministry of Health and Social Assistance, aims to better characterize the epidemiology and transmission of AFI pathogens including SARS-CoV-2. Patients ≥ 2 years of age who presented to study sites with an undifferentiated fever either measured (≥ 38.0 °C) or by history, or with new onset anosmia or ageusia were invited to participate. Study staff (all medical doctors) performed enrollment five days per week from 8am to 5pm. Questionnaires were administered using the KoBo Toolbox data collection platform (www.kobotoolbox.org) on electronic tablets to collect individual-level covariates including demographic data (age, gender, race-ethnicity); comorbid medical conditions (hypertension, coronary heart disease, diabetes, active cancer, chronic kidney disease, stroke, asthma, chronic obstructive pulmonary disease); weight and height; primary occupation; symptom onset date; and number, date, and type of COVID-19 vaccines received. Nasopharyngeal swabs and venous blood were collected from all participants at the time of enrollment. Blood was processed as sera and biological samples were stored at -80°C.

To assess the association between peri-infection anti-S levels and risk of SARS-CoV-2 infection we used a case-negative approach that first assigned study participants into two groups based on SARS-CoV-2 virological test result. We then assessed crude anti-S levels between groups and subsequently performed univariable and multivariable binomial logistic regression with anti-S levels categorized by quartile. Anti-S antibody levels at the time of SARS-CoV-2 diagnosis (peri-infection) were considered to reflect antibody levels at the time of infection.

Written consent was obtained for all participants. For children <18 years old, except emancipated minors, consent was obtained from the legal guardian. Written assent was provided by adolescents 14-17 years old, and verbal assent by children 7-13 years old. The study was reviewed and approved by the National Council of Bioethics in Health, Santo Domingo (013-2019), the Institutional Review Board of Pedro Henríquez Ureña National University, Santo Domingo, and the Mass General Brigham Human Research Committee, Boston, USA (2019P000094). Study procedures and reporting adhere to STROBE criteria for observational studies.

### Immunoassay characteristics

Serum pan-immunoglobulin antibodies against the SARS-CoV-2 spike (anti-S) glycoprotein were measured at the Brigham and Women’s Hospital, Boston, USA, on the Roche Elecsys SARS-CoV-2 electrochemiluminescence immunoassay that uses a recombinant protein modified double-antigen sandwich format (Roche Diagnostics, Indianapolis, USA). The assay was calibrated with positive and negative quality controls before analyses. Values were quantified between 0.40 and 250 U/mL representing the primary measurement range, with values below 0.40 U/mL reported as 0.40 U/mL. Samples with measured values >250 U/mL underwent automated 1:50 dilution with further 1:10 dilution for samples >12,500 U/mL, representing an upper limit of detection of 125,000 U/mL. Samples were considered reactive according to the manufacturer cutoff index (COI) (≥ 0.8 U/mL). Values are reported as binding antibody units (BAU), that equal Elecsys SARS-CoV-2 anti-S U/mL in accordance with manufacturer’s recommendations and the WHO International Standard and International Reference Panel for anti-SARS-CoV-2 immunoglobulin.^14^ Assay performance measures reported by the largest non-manufacture-sponsored study registered a specificity of 99.8% (CI 99.3-100) and sensitivity of 98.2% (CI 96.5-99.2).^15^

### Virological assays

Acute SARS-CoV-2 infection was assessed using real-time reverse transcriptase polymerase chain reaction nucleic acid amplification tests (NAAT) on nasopharyngeal specimens using the Allplex SARS-CoV-2 kit (Seegene, Seoul, South Korea) that amplifies the E, N and the RdRP genes. Conditions for amplifications were 50 °C for 20 min, 95 °C for 15 min, followed by 45 cycles of 95 °C for 10 s and 60 °C for 15 s and 72 °C for 10 s. Samples were considered positive according to the manufacturer recommendations with a cycle threshold value of less than 37. A cycle threshold value of ≥38 or more was defined as a negative test. Genomic sequencing was performed on a subset of NAAT-positive samples (**Supplementary Methods**).

### Classification and statistical analysis

Mean SARS-CoV-2 seroprevalence and number of COVID-19 vaccines received were analyzed by seven-day intervals, starting on the first day of the study period. We defined viral strain transmission phases according to the predominant circulating viral strain based on genome sequencing of 237 SARS-CoV-2 RT-PCR positive study samples: 22 March 2021 to 15 August 2021 (pre-Delta), 16 August to 23 December 2021 (Delta), 24 December 2021 to 30 April 2022 (BA.1, Omicron), and 1 May to 17 August 2022 (post-BA.1). Because phases varied in duration, we created a second date partition that captured largely similar three-to-four-month time intervals: Mar-June 2021 (22 March to 30 June 2021), July-September 2021, October-December 2021, January-April 2022, and May-August 2022 (1 May to 17 August 2022). Data were analyzed by date of participant enrollment. Days post-symptom onset (DPSO) was calculated by subtracting the symptom-onset date from the date of enrollment. For 10 participants without symptom onset date, DPSO was imputed as the DPSO mode for all other participants. Age was aggregated into three groups, 2–17, 18–54, and ≥ 55 years, with cutoffs selected to capture groups with documented differences in seroprevalence in the Dominican Republic, while minimizing data sparsity among older adults. Given this was an observational study of prospectively enrolled patients, all eligible participants with required data were included and no sample size power calculation was performed.

Analyses and data visualization were performed using the R statistical programming language (R version 4.1.3, 2022-03-10), with finalfit (glm) for univariable and multivariable logistic regression, and visreg and ggplot2 for data visualization.^16–18^

### Data sources

National SARS-CoV-2 cases, deaths, and vaccination data were obtained from COVID-19 GitHub repository.^6^ Other data were enumerated during the study.

## Results

Between 22 March 2021 and 17 August 2022, 2,814 eligible patients were invited to participate of which 2,502 (89.0%) were enrolled. Of these, 2,300/2,502 (91.9%) had complete virological, serological and demographic data and are included in analyses (**Supplementary Fig 1**). The median age was 31 years (IQR 7, 55) and 1,422/2,300 (61.8%) were female. The mean interval between symptom onset and enrollment was 4.0 days (Gmd 2.5) for all participants and 3.7 days (Gmd 2.1) for SARS-CoV-2 NAAT positive participants (**Supplementary table 1**). Overall SARS-CoV-2 NAAT test positivity was 22.4% (517/2,300) with trends over time detailed in **Fig 1. Table 1** presents the characteristics of study participants by SARS-CoV-2 NAAT status.

**Table 1.**
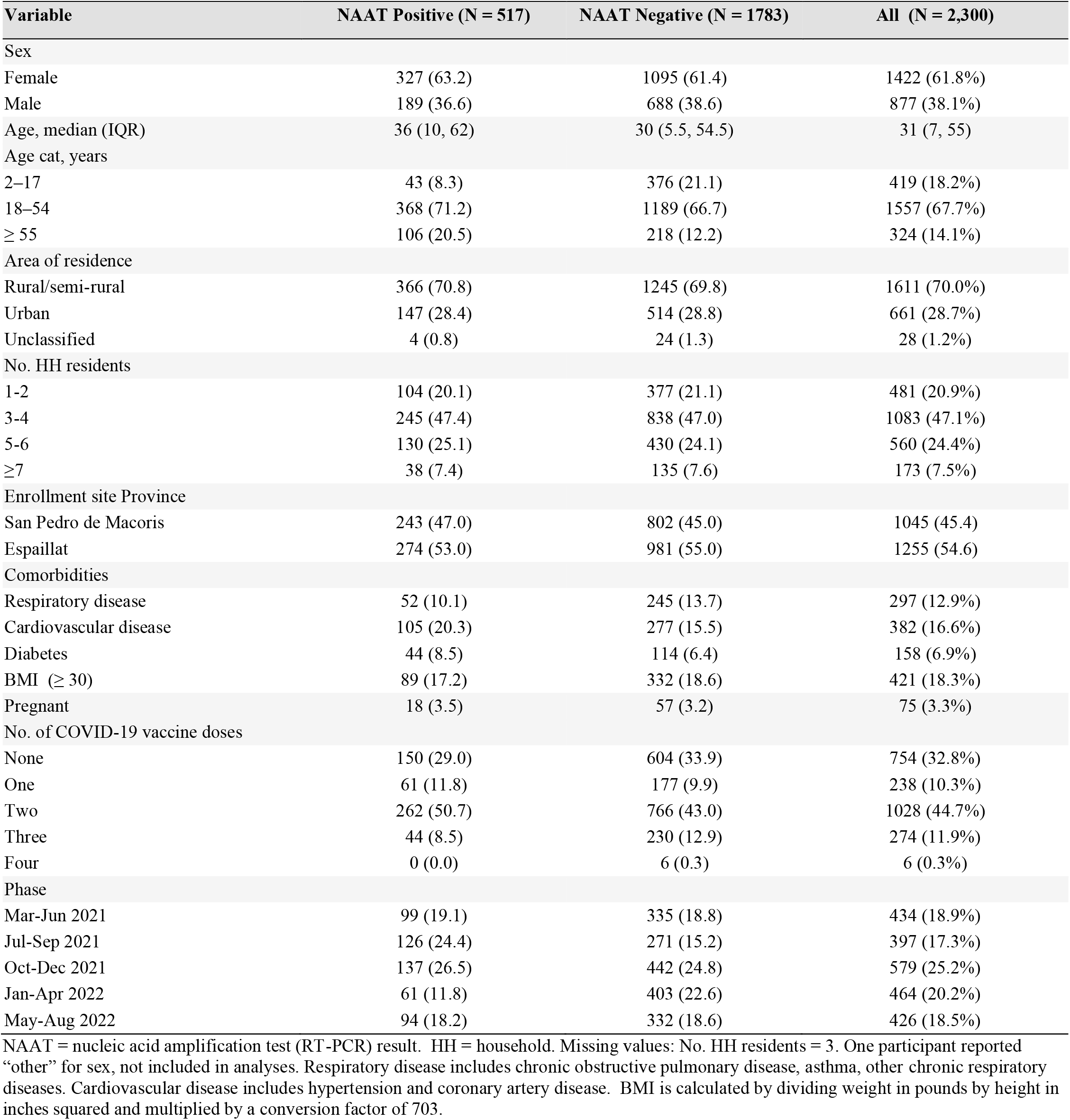
Population characteristics of study participants by SARS-CoV-2 NAAT test status — Dominican Republic, March 2021–August 2022.

**Figure 1.**
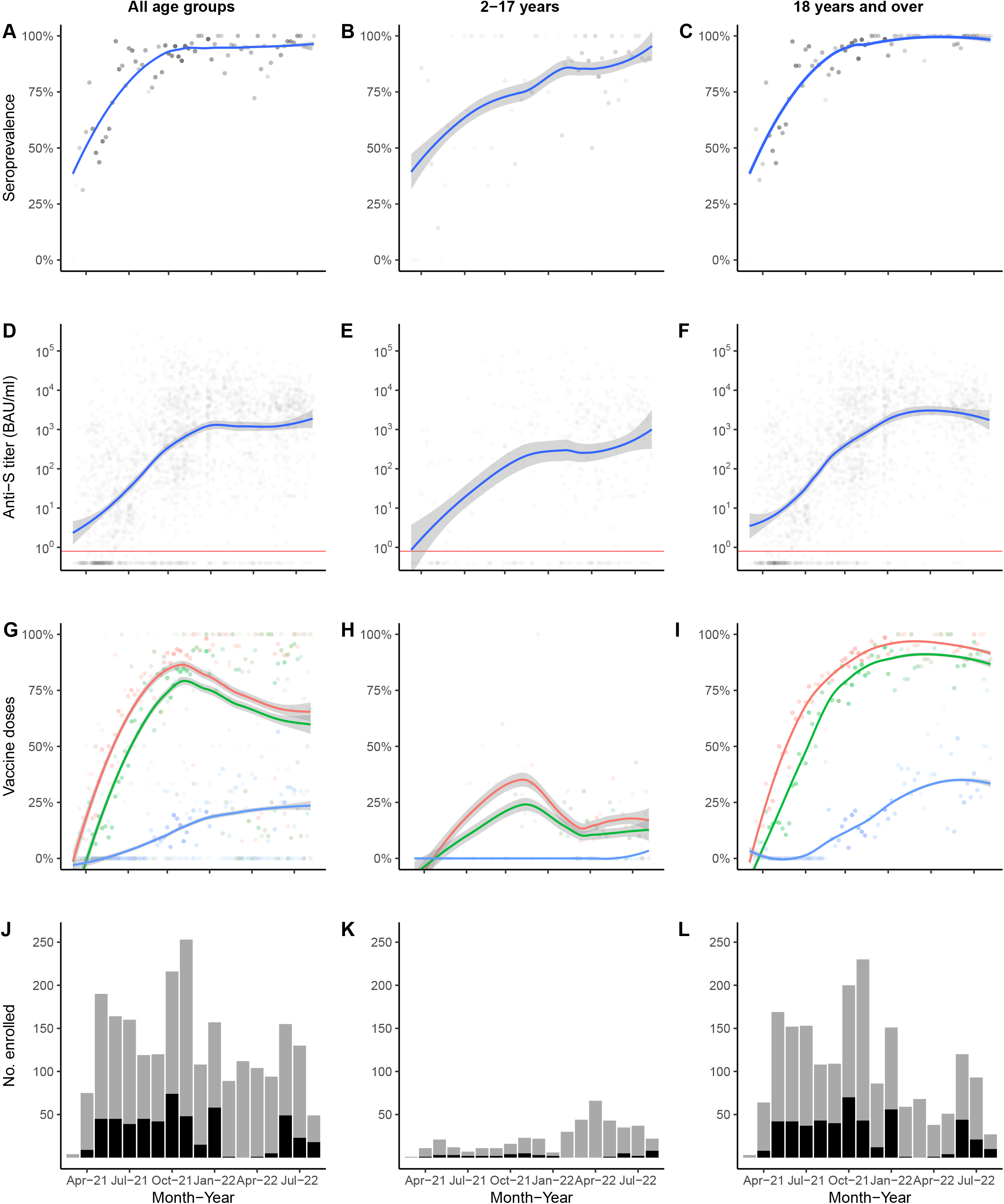
SARS-CoV-2 anti-spike seroprevalence, titers, vaccine doses and participant enrollment by age group, Dominican Republic, March 2021–May 2022 (N=2,300). (A–C) Anti-S seroprevalence among study participants. Gray dots indicate the weekly mean values, with increased dot intensity reflecting more observations. Blue line indicates the LOESS smoothed seroprevalence, with gray shading indicating the 95% confidence interval (95% CI) around the smoothed estimate. (D–F) Anti-S antibody titers by week plotted on a log scale with each gray dot indicating a unique study participant (n=1,910), blue lines, and gray shading reflecting LOESS smoothed anti-S levels and associated 95% CI. Horizontal red line indicates the manufacturer recommended cutoff index (COI 0.800 BAU/ml) with values above the line representing a positive result and values below the line a negative result. (G–I) Fraction of weekly enrolled participants who had received one or more (red dots), two or more (green dots), or three (blue dots) COVID-19 vaccine doses, with increased dot intensity reflecting more observations. Coloured lines indicate the LOESS smoothed fraction, with gray shading indicating the 95% CI. (J–L) Number of study participants enrolled per month, with bars representing total monthly enrollment, gray shading indicating SARS-CoV-2 NAAT negative, and black shading representing NAAT positive participants. Labels on the x-axis indicate complete months except March 2021, which represents participants enrolled from 22 March 2021, and August 2022, which represents enrollment through 17 August 2022.

### Changes in COVID-19 vaccination rates

Following the launch of the national COVID-19 vaccination campaign in late February 2021, coverage among the study population increased rapidly, consistent with nationally reported data. By December 2021 about 75% of study participants had completed a two-dose primary series (**Fig 1**). Unexpectedly, overall vaccination rates declined after December 2021, a finding attributable to an increase in younger pediatric patients who were ineligible for COVID-19 vaccines at the time, a finding consistent across study sites (**Supplementary Fig 2 & 3)**. Vaccination coverage among adults remained high through the remainder of the study and by June–August 2022 about 80% of adults had completed a primary vaccine series and about 35% had received a third vaccine dose. Vaccination coverage among adult study participants appeared to be modestly higher than national reported COVID-19 data,^7^ but without age-stratified national vaccination data, which were not available, direct comparisons could not be made.

### Temporal changes in anti-S seropositivity and levels

Between the March–June 2021 and May–August 2022 study periods, the proportion of participants testing positive for anti-S antibodies increased from 61.1% to 95.8% and GMT and median titer values increased 202- and 757-fold respectively (**Fig 2 A, Table 2)**, with near-log linear increases across the study population through January 2022 when overall GMT flattened. Figure 2 demonstrates the trend in overall antibody titers during the study period (A) and further stratified by age group (B) and vaccination status (C). We observed progressive increases in anti-S titers over time across all age groups and, interestingly, even within each vaccine dose category. For example, among recipients of two vaccine doses, GMT increased from 72.1 BAU/mL (40.1, 129.7) during the March–June 2021 period to 2153.2 BAU (1684.7, 2752.1) during the May–August 2022 study period, with similar trends across recipients of one vaccine dose (**Fig 2C, Supplementary table 3)**. A less pronounced increase was observed across three vaccine dose recipients, which demonstrated high titers, measured on a logarithmic scale, across all time periods. Increases in GMT over time within vaccine dose categories likely represents ongoing immunological exposure due to SARS-CoV-2 infections and transition from the less immunogenic Sinovac vaccine early in the national vaccination campaign to the mRNA BNT162b2 (Pfizer/BioNTech) vaccine in late 2021. As evidenced by progressively increasing anti-S titers over time among unvaccinated study participants, and consistent with nationally reported data, substantial SARS-CoV-2 transmission continued through most of the study period. Yet, despite ongoing transmission, anti-S titers remained substantially lower in unvaccinated than vaccinated participants. For example, during May-August 2022, anti-S GMTs among unvaccinated participants represented 25.7%, 13.1%, and 6.3% of anti-S GMTs observed among recipients of one, two and three dose vaccine doses, respectively (**Fig 2C, Supplementary table 3)**.

**Table 2.** SARS-CoV-2 anti-S antibody serostatus, geometric mean, and median titers by study period interval — Dominican Republic, March 2021 – August 2022.

| Study period | N | Seropositive, n (%) | GMT (95% CI) | Median titer, BAU/ml (Q1, Q3) |
| --- | --- | --- | --- | --- |
| Mar-Jun 2021 | 434 | 265 (61.1) | 6.6 (5.1, 8.7) | 3.8 (0.4, 57.5) |
| Jul-Sep 2021 | 397 | 344 (86.6) | 62.8 (45.8, 86.0) | 62.5 (6, 581.8) |
| Oct-Dec 2021 | 579 | 543 (93.8) | 559.4 (439.8, 711.5) | 781.7 (104.9, 4813.5) |
| Jan-Apr 2022 | 463 | 434 (93.7) | 1180.3 (906.3, 1537.2) | 2578 (390.8, 8137.5) |
| May-Aug 2022 | 427 | 409 (95.8) | 1332.4 (1055.3, 1682.3) | 2876 (775.8, 5483.5) |

GMT, geometric mean titer. Study periods indicate complete months except March 2021, which represents participants enrolled from 22 March 2021, and August 2022, which represents enrollment through 17 August 2022. Seropositive, anti-spike antibodies above the manufacturer cutoff index ( $\geq 0.8$ BAU/ml). N= 2,300.

**Figure 2.**
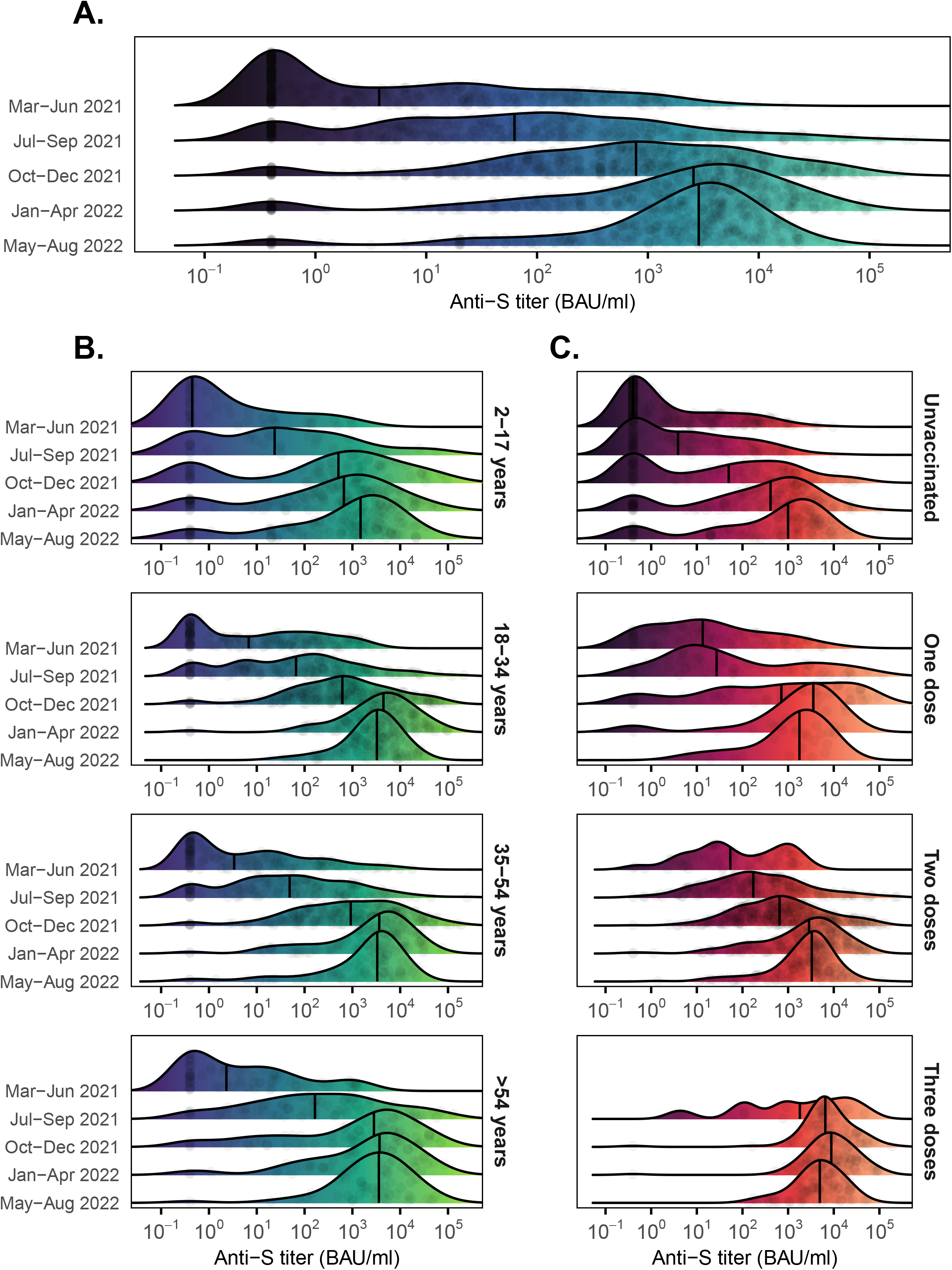
Distribution of SARS-CoV-2 anti-spike titers among study participants, Dominican Republic, March 2021 – August 2022. (A) Smoothed density plot demonstrates log adjusted distribution of anti-S antibody titers (x-axis) among all study participants with y-axis indicating date interval when study participants were enrolled from earliest (March–June 2021, upper) to latest (May – August 2022, lower) (n=2,300). Study interval labels on the y-axis (left) indicate complete months except March 2021, which represents participants enrolled from 22 March 2021, and August 2022, which represents enrollment through 17 August 2022. (B) Similar smoothed density plots with study participants stratified by age group (n=2,300). Dark purple shading indicates lower anti-S titers and light green higher titers. (C) Darker red shading indicates lower anti-S titers and light orange higher titers. Data stratified by number of COVID-19 vaccine doses received from none (unvaccinated, top plot) to three (bottom plot) (n=2,293). Six participants that received four COVID-19 vaccine doses not included. Values for Mar–Jun 2021-three vaccine dose plot not shown given sparsity of data points (n=1). For all plots, gray circles represent jitter adjusted individual study participant values. Median values indicated with a narrow vertical black line. Lower limit of assay measurement is 0.4 BAU/mL and values <0.4 BAU/mL are represented as 0.4 BAU/mL, with smoothing extending curves below the lower measurement limit. Therefore, density plot shading is used for illustrative purposes with summarized data used for plots in Table 2 and Supplementary Tables 2 and 3.

### Association between anti-S titers and SARS-CoV-2 NAAT status

The increase in anti-S levels across all demographics during the study period was striking, yet the implications for immunological protection was unclear. Therefore, we used a case negative approach to assess if simple unadjusted anti-S levels were associated with NAAT test result. We identified a consistent inverse association across all phases of transmission (**Table 3**). Using multivariable analyses, we again identified an inverse association between anti-S quartile and odds ratio for a positive NAAT, with results demonstrating a clear biological gradient (**Table 4**). Younger and older age were associated with lower (0.45 (0.29–0.68, p<0.001)) and higher (1.58 (1.19–2.07, p=0.001) odds ratios for a positive NAAT test (2–17 years and ≥ 55 years versus 18–54 years, respectively) across all phases of transmission combined, and similar but largely non-significant trends when stratified by phase (**Supplementary tables 4–8**). No consistent association was observed between NAAT status and sex or number of COVID-19 vaccine doses received after controlling for anti-S levels (**Supplementary tables 4–8**).

**Table 3.** Geometric mean and median anti-S antibody titers by SARS-CoV-2 NAAT status and phase of predominant circulating viral strain, March 2021 – August 2022.

| Phase | SARS-COV-2 NAAT | N (%) | GMT, BAU/ml (95% CI) | Fold difference (GMT) | Median titer, BAU/ml (Q1, Q3) | Fold difference (Median titer) |
| --- | --- | --- | --- | --- | --- | --- |
| **Pre-Delta** | Neg | 495 (76.6) | 14.1 (10.9, 18.2) | 3.4* | 13.3 (0.8, 132.8) | 5.5 |
|  | Pos | 151 (23.4) | 4.1 (2.6, 6.5) |  | 2.4 (0.4, 24.4) |  |
| **Delta** | Neg | 553 (72.4) | 604.8 (475.3, 769.7) | 3.9* | 792.3 (127.8, 4805) | 4.5 |
|  | Pos | 211 (27.6) | 154.7 (97.8, 244.7) |  | 176.2 (24.7, 1638.5) |  |
| **Omicron (BA.1)** | Neg | 403 (86.9) | 1288.3 (965.4, 1719.2) | 1.8* | 2822 (511.1, 8656) | 3.4 |
|  | Pos | 61 (13.1) | 713.8 (375.1, 1358.5) |  | 837.2 (126.8, 5739) |  |
| **Omicron (BA.2/4/5)** | Neg | 332 (77.9) | 1541.4 (1183.4, 2007.6) | 2.0* | 3202 (1011, 6173) | 1.7 |
|  | Pos | 94 (22.1) | 759.6 (468.1, 1232.6) |  | 1835 (197.7, 3882.2) |  |
| **Total** | Neg | 1783 (77.5) | 300.6 (256.5, 352.4) | 3.5* | 725 (31.6, 4351.5) | 6.0 |
|  | Pos | 517 (22.5) | 85.8 (62.9, 117.1) |  | 121.2 (4.6, 1905) |  |

NAAT = nucleic acid amplification test (RT-PCR) result. GMT = geometric mean titer. Fold difference is the GMT or median titer of NAAT negative participants divided by the titer of NAAT positive participants. Phases based on dominant circulating strain: 22 March 2021 to 15 August 2021 (pre-Delta, primarily Alpha and Mu), 16 August to 23 December 2021 (Delta), 24 December 2021 to 30 April 2022 (Omicron, BA.1), and 1 May to 17 August 2022 (Omicron, BA.2, BA.4, BA.5). T-test p-value for GMT difference between NAAT negative and positive study participants: \* = $p$ -value $<0.001$ .

**Table 4.** Multivariable odds ratios for a SARS-CoV-2 NAAT positive test result by phase of predominant circulating viral strain, March 2021 - August 2022.

| Anti-S titer, ... | All (N=2,300) | Pre-Delta (N=646) | Delta (N=764) | Omicron BA.1 (N=464) | Omicron BA.2, BA.4, BA.5 (N=426) |
| --- | --- | --- | --- | --- | --- |
| Q1 | Ref | Ref | Ref | Ref | Ref |
| Q2 | 0.55 (0.40-0.74)*** | 0.25 (0.14-0.44)*** | 0.38 (0.23-0.62)*** | 0.69 (0.27-1.76) | 0.60 (0.30-1.17) |
| Q3 | 0.38 (0.27-0.55)*** | 0.13 (0.07-0.25)*** | 0.31 (0.19-0.51)*** | 0.46 (0.16-1.27) | 0.30 (0.14-0.60)** |
| Q4 | 0.27 (0.18-0.40)*** | 0.13 (0.06-0.25)*** | 0.31 (0.18-0.54)*** | 0.14 (0.04-0.42)** | 0.22 (0.09-0.50)*** |

Lambda strains), 16 August to 23 December 2021 (Delta), 24 December 2021 to 30 April 2022 (Omicron, BA.1), and 1 May to 17 August 2022 (Omicron, BA.2, BA.4, BA.5).

## Discussion

We report on the temporal change in SARS-CoV-2 anti-S antibody prevalence and levels over 18 months among patients enrolled through a longitudinal acute febrile illness surveillance platform in the Dominican Republic. The study period aligned with the beginning of the national COVID-19 vaccination campaign in late February 2021, providing a unique opportunity to characterize the evolution of anti-SARS-CoV-2 antibodies in this setting and across a population that was largely COVID-19 vaccine-naive early in the study period. We observed a progressive increase in seroprevalence of SARS-CoV-2 anti-S antibodies — reflecting vaccination, infection, or both — increasing from 61% to 96%. Strikingly, geometric mean and median titers increased about 200- and 760-fold respectively during the study period. To understand the implications of these findings for public health we used a case-negative approach to assess antibody levels between NAAT positive vs negative cases. We identified a consistent inverse association between anti-S titers and odds of testing positive for SARS-CoV-2 by NAAT and extended those findings to phases of predominantly pre-Delta, Delta, Omicron-BA.1, and Omicron derivative strain waves of transmission.

Anti-S levels were lower amongst those who tested positive vs negative for acute SARS-CoV-2 infection, a trend that was consistent across strains and after adjustment for potential confounders. The likelihood of testing positive was reduced by about 45%, 60% and 75% across the second, third and fourth anti-S titer quartiles when compared to the first quartile. This finding aligns with several correlates of protection studies that reports that anti-spike binding antibodies track with risk of SARS-CoV-2 infection.^5–7,19^ We built on the findings from these prior studies, that were conducted prior to widespread transmission of variants of concern, to include transmission waves that were primarily driven by Delta, Omicron-BA.1 and subsequent Omicron strains (BA.2/BA.4/BA.5), and document similar predictive utility of anti-S antibodies against these strains. These findings suggest that binding antibodies, at least total anti-S antibodies as measured in this study, track with functional measures of immunological protection such as viral neutralization, Fc-function, and potentially T cell responses, as previously reported.^19–21^ Our findings suggest that while likely inappropriate for adjudicating vaccine efficacy and vaccine approval, total SARS-CoV-2 binding antibodies are reasonable surrogate markers of immunological protection against infection, including infection by emerging strains with substantial immune-evasion capacity.

Given the relative simplicity, high throughput capacity, and cost-effectiveness of measuring anti-S antibodies versus live or pseudoviral neutralizing activity, this approach may be suitable for characterizing population level immunological protection, parameterizing transmission and prediction models and, in turn, informing national and regional public health policy.

For our analyses we assumed *a priori* that anti-S levels measured at the time that patients seek care is a reasonable surrogate measure of anti-S levels at the time of infection, including NAAT-positive cases, an approach that while not well characterized has been previously described.^22^ Most of the SARS-CoV-2 NAAT-positive cases can be assumed to have been mounting a humoral response to the acute infection at the time of sample collection, with individuals previously exposed to SARS-CoV-2 antigens expected to mount a more rapid and robust anamnestic response. Yet whether this would obscure differences in antibody levels between groups, if present, was unclear prior to the study. Although this was not the case, observed differences in anti-S levels by NAAT status were likely attenuated. To probe this question, we assessed if antibody levels trended higher based on the number of days between symptom onset and sample collection but were unable to detect a clear trend including after stratifying by number of vaccine doses received (**Supplementary Fig 4**), potentially because 50% of NAAT-positive-cases were collected within three days of symptom onset, and 90% within six days (**Supplementary table 1)**. We also performed sensitivity analyses comparing samples collected 0–4 versus ≥5 DPSO and identified broadly similar findings, although the biological gradient observed for samples collected 0–4 DPSO was less clearly defined for samples collected ≥5 DPSO (**Supplementary table 9**).

This study has multiple strengths. Dedicated study staff prospectively enrolled study participants using well defined procedures, administered standardized survey questionnaires, and simultaneously collected respiratory and blood samples. Enrollment of eligible patients was high for this type of surveillance study (89%). Sera were tested with a widely used and validated immunoassay and antibody titers were reported as internationally standardized units so our findings can be compared across other settings. By building off other work, we developed a novel methodological approach to understand temporal changes in population level SARS-CoV-2 antibodies, methods that may be applicable to other settings. We performed genomic sequencing of a relatively large number of samples from among the current study population, and therefore were able to characterize timing of predominant SARS-CoV-2 strain transmission. Lastly, we enrolled participants across geographically discrete settings, limiting the potential for study-site specific biases, with findings consistent across sites (**Supplementary Fig 2-3**).

Yet there were at least five limitations. First, about 8% of enrolled study participants did not have serology or NAAT data and were excluded from analyses, but the demographic profile of these individuals largely reflected the final study population. Second, demographic, comorbidities, and COVID-19 vaccination data were self-reported, which may introduce recall or social-desirability biases, potentially impacting our findings in either direction. Third, we used a total anti-S antibody immunoassay and findings may be different for other assays that measure binding or neutralizing antibodies. Fourth, the sensitivity of NAAT for the detection of acute symptomatic SARS-CoV-2 infection is estimated to be between 70% and 95%,^23^ and therefore some infections may have been mis-classified as non-infections, which would attenuate the differences in anti-S levels between cases and non-cases reported in this study. Fifth, the study was conducted among a discrete population of patients seeking healthcare for undifferentiated fever and therefore changes in antibody levels may not reflect the broader population, which would limit generalizability. However, as described above, findings were similar across our two geographically discrete study sites, suggesting findings may be comparable across similar healthcare settings in the country.

In summary, we believe there are three broad findings from this study. First, we provide the first documentation of longitudinal changes in SARS-CoV-2 antibody titers following the launch of a national vaccination campaign. Second, we document that total anti-spike binding antibody levels track closely with risk of infection across multiple viral strains, including strains with highly effective immune evasion capacity. And third, we present what we believe is a novel approach to monitoring changes in immune biomarkers among discrete populations, an approach that is relatively simple and can leverage existing surveillance infrastructure. Given future SARS-CoV-2 variants of concern and other emerging pathogens will occur, establishing pragmatic, sustainable and real time methods to estimate population immune markers while simultaneously assessing strain-specific risks of infection may prove a valuable complement to existing surveillance infrastructure.

Disclaimer: The findings and conclusions in this report are those of the authors and do not necessarily represent the official position of the U.S. Centers for Disease Control and Prevention

## Data Availability

All data produced in the present study are available upon reasonable request to the authors.

## Acknowledgments

We would like to thank the many participants that volunteered to participate in this study. We would also like to thank the study staff that collected the field data, the Dominican Republic Ministry of Health and Social Assistance and the Pedro Henriquez Ureña National University for their commitment and support for the study.

