## Supplementary for "Integrated SARS-CoV-2 serological and virological screening across an acute fever surveillance platform to monitor temporal changes in anti-spike antibody levels and risk of infection during sequential waves of variant transmission — Dominican Republic, March 2021 to August 2022"

### Supplementary Methods

#### Viral genome sequencing and analysis

A total of 201 study samples were sequenced with analysis performed using the ARTIC pipeline and Oxford Nanopore Technologies tools.[1] Samples were randomly selected from NAAT positive samples from April 2021 to February 2022. Total nucleic acids (TNA) were extracted from nasopharyngeal samples using Zymo Research Quick DNA/RNA pathogen MiniPrep (Zymo Research Corp. Irvine, CA). Samples were initially screened by RT-PCR targeting the Sars-CoV-2 E gene as previously described.[2] Any sample with a Ct-value ≤25 was sequenced to determine Sars-CoV-2 variant. SARS-CoV-2 genome amplification was performed on the TNA samples using the NEBNext® ARTIC SARS-CoV-2 Companion Kit for Oxford Nanopore Technologies (New England Biolabs, Ipswich, USA) using the ARTIC V3 primers following manufacturer's instructions. The amplicons were sequenced using the SQK-LSK109 and EXP-NBD196 library prep kits with the Mk1B sequencer and R9.4.1 flow cell (Oxford Nanopore Technologies, Oxford, UK) following manufacturer's protocols. The sequence data were basecalled using Guppy v6.1.5 with the high accuracy (HAC) model with barcodes required on both ends of the amplicons. The data were analyzed using the EPI2ME labs ARTIC workflow v0.3.18 (Oxford Nanopore Technologies, Oxford, UK) with default settings, which used Nextclade v1.11.0, and Pangolin v4.1.1 for clade and lineage classification.[3–5] Samples which were missing more than three kilobases of genome sequence were not considered for analysis.

An additional 36 samples were sequenced using the Illumina DRAGEN COVID Lineage 3.5.11. Samples were randomly selected from NAAT positive samples from March to August 2022. TNA were extracted from nasopharyngeal samples using automatic magnetic extraction method with the RADI EXTRACTOR 192 and the RADI 192 viral DNA/RNA extraction kit (KH Medical, South Korea). Samples were screened using real-time reverse transcriptase polymerase chain reaction nucleic acid amplification tests (NAAT) of nasopharyngeal specimens using the Allplex SARS- CoV-2 kit (Seegene, Seoul, South Korea) that amplifies the E, N and the RdRP genes. Samples with a Ct-value ≤25 were sequenced. Library preparation was performed using the Illumina Covid Seq assay with the ARTIC V3 primers and were sequenced with Illumina´s MiSeq reagent kit V2, 300 cycles according to manufacturer specifications. Sequencing data was analyzed with the DRAGEN COVID Lineage 3.5.11 application on Illumina´s Basepace sequence hub with the default settings which used Nextclade 2.6.0 and Pangolin 4.1.2 pangolin-data 1.14 for clade and lineage classification.

### Supplementary results

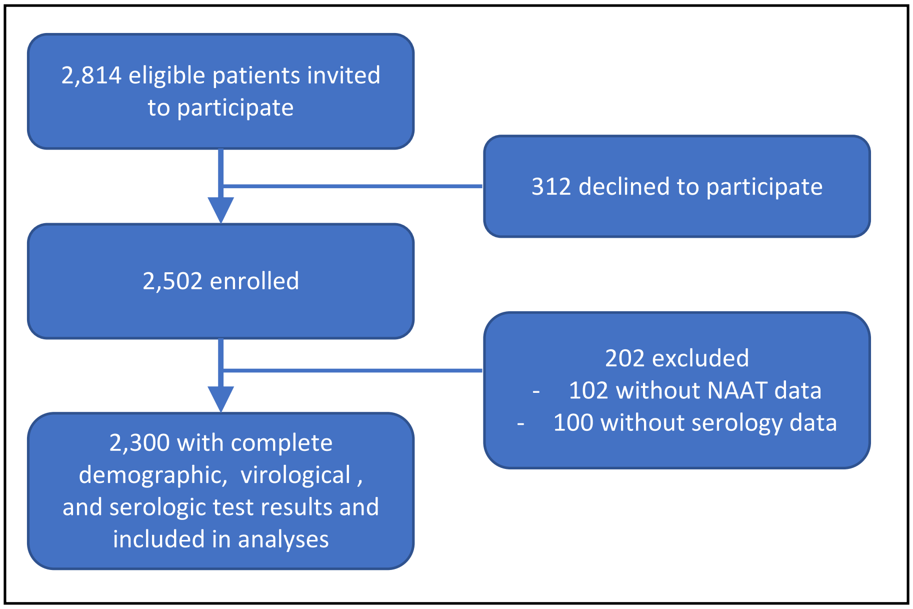


**Supplementary Figure 1. Study participant enrollment**

**Supplementary Table 1. Days from symptom onset to enrollment by SARS-CoV-2 NAAT status**

| NAAT Status | n | Mean, days | Gmd | 25% | 50% | 75% | 90% | 95% | Range, days |
|---|---|---|---|---|---|---|---|---|---|
| Positive | 517 | 3.7 | 2.1 | 2 | 3 | 5 | 6 | 7 | 1–14 |
| Negative | 1783 | 4.1 | 2.6 | 2 | 4 | 5 | 7 | 8 | 0–20 |
| All | 2300 | 4 | 2.5 | 2 | 4 | 5 | 7 | 8 | 0–20 |

Blood collected for anti-S serological tested at the time of enrollment. Gmd = Gini's mean difference. Percent represents the 25$^{th}$, 50$^{th}$, 75$^{th}$, 90$^{th}$, and 95$^{th}$ quantiles, with the 50$^{th}$ quantile representing the median number of days from symptom onset to enrollment/sample collection.

**Supplementary table 2. Geometric mean and median anti-S antibody titers by age group and study period, Dominican Republic, March 2021 – August 2022**

| Age, | Study period | N | Geometric mean (95% CI) | Median titer, BAU/ml (Q1, Q3) |
|---|---|---|---|---|
| 2–17 | Mar-Jun 2021 | 46 | 2.7 (1.3, 5.8) | 0.4 (0.4, 14.5) |
| | Jul-Sep 2021 | 28 | 29.3 (7.6, 112.9) | 24.4 (0.8, 254.6) |
| | Oct-Dec 2021 | 61 | 193.7 (69.6, 539.5) | 510.4 (11.6, 3548) |
| | Jan-Apr 2022 | 146 | 260.4 (149.1, 454.7) | 663.6 (43.2, 2743) |
| | May-Aug 2022 | 138 | 481.6 (281.8, 822.9) | 1481.5 (96.8, 4115) |
| 18-54 | Mar-Jun 2021 | 315 | 8.1 (5.8, 11.1) | 5.6 (0.4, 80.2) |
| | Jul-Sep 2021 | 297 | 51.9 (36.5, 73.9) | 60.2 (5.4, 475.5) |
| | Oct-Dec 2021 | 450 | 613.2 (478.2, 786.3) | 690 (114.2, 4307.5) |
| | Jan-Apr 2022 | 259 | 2598.2 (1983.4, 3403.6) | 4084 (1250.5, 9794.5) |
| | May-Aug 2022 | 236 | 2105.1 (1673.9, 2647.2) | 3286.5 (1234.2, 5793.5) |
| ≥55 | Mar-Jun 2021 | 73 | 5 (2.7, 9.4) | 2.3 (0.4, 22.6) |
| | Jul-Sep 2021 | 72 | 185.9 (86.4, 399.7) | 164.8 (19.8, 1615.8) |
| | Oct-Dec 2021 | 68 | 788.6 (351.9, 1767.6) | 2824 (78.3, 7154.8) |
| | Jan-Apr 2022 | 58 | 1563.8 (762.4, 3207.7) | 3694.5 (346.5, 8733.5) |
| | May-Aug 2022 | 53 | 2460.4 (1415.9, 4275.4) | 3579 (1019, 6951) |

GMT = geometric mean titer. Study periods indicate complete months except March 2021, which represents participants enrolled from 22 March 2021, and August 2022, which represents enrollment through 17 August 2022.

**Supplementary table 3. Geometric mean and median anti-S antibody titers by number of vaccine doses and study period, Dominican Republic, March 2021 - August 2022**

| No. COVID-19 vaccine doses | Study period | N | Geometric mean (95% CI) | Median titer, BAU/ml (Q1, Q3) |
|---|---|---|---|---|
| None | Mar-Jun 2021 | 281 | 2.8 (2.1, 3.8) | 0.4 (0.4, 20.6) |
| | Jul-Sep 2021 | 118 | 5.5 (3.4, 8.9) | 3.9 (0.4, 37.5) |
| | Oct-Dec 2021 | 87 | 27.9 (13, 59.8) | 49.9 (0.4, 606) |
| | Jan-Apr 2022 | 139 | 153.2 (88.2, 266.3) | 411.8 (34.9, 1585) |
| | May-Aug 2022 | 129 | 282.5 (160.7, 496.6) | 1000 (52, 3132) |
| One | Mar-Jun 2021 | 89 | 17.9 (10, 32.2) | 13.7 (1.8, 115) |
| | Jul-Sep 2021 | 55 | 62.8 (24.8, 159.1) | 27.1 (5.4, 1125.1) |
| | Oct-Dec 2021 | 48 | 399.6 (135.6, 1177.9) | 712.4 (33.4, 7489.8) |
| | Jan-Apr 2022 | 26 | 1216 (395.9, 3734.7) | 3565.5 (785.4, 5005) |
| | May-Aug 2022 | 22 | 1099.8 (458.7, 2636.7) | 1784.5 (701.3, 4120) |
| Two | Mar-Jun 2021 | 64 | 72.1 (40.1, 129.7) | 56.4 (15.4, 631.4) |
| | Jul-Sep 2021 | 214 | 209.4 (148.6, 295.1) | 172.6 (38.1, 1051.5) |
| | Oct-Dec 2021 | 367 | 703.6 (560.9, 882.6) | 649.2 (153.8, 2857) |
| | Jan-Apr 2022 | 202 | 1891.1 (1394.9, 2563.7) | 2877 (666.6, 8203.2) |
| | May-Aug 2022 | 180 | 2153.2 (1684.7, 2752.1) | 3302.5 (1225.5, 5419.2) |
| Three | Mar-Jun 2021 | 0 | *NC* | *NC* |
| | Jul-Sep 2021 | 10 | 1277.8 (161.6, 10100.2) | 2231.1 (277.6, 13452.2) |
| | Oct-Dec 2021 | 76 | 6844.5 (4775.2, 9810.6) | 6532.5 (4523.5, 11250.5) |
| | Jan-Apr 2022 | 95 | 8209.3 (6044.9, 11148.6) | 8844 (4483, 19871.5) |
| | May-Aug 2022 | 92 | 4485.5 (3477.5, 5785.7) | 4950 (2290, 10284.5) |
| Four | Mar-Jun 2021 | 0 | *NC* | *NC* |
| | Jul-Sep 2021 | 0 | *NC* | *NC* |
| | Oct-Dec 2021 | 1 | 7782 (*NC*, *NC*) | 7782 (*NC, NC)* |
| | Jan-Apr 2022 | 1 | 41355 (*NC*, *NC*) | 41355 (*NC*, *NC*) |
| | May-Aug 2022 | 4 | 6377.9 (1951.7, 20841.9) | 6589 (5293.2, 9048) |

GMT = geometric mean titer. NC = not calculated. Study periods indicate complete months except March 2021, which represents participants enrolled from 22 March 2021, and August 2022, which represents enrollment through 17 August 2022.

**Supplementary Figure 2. Anti-spike seroprevalence, titers, vaccine doses and participant enrollment by age group, Hospital Dr. Antonio Musa, San Pedro de Macoris, Dominican Republic, April 2021–August 2022 (n=1,045)**

**Supplementary Figure 3. Anti-spike seroprevalence, titers, vaccine doses and participant enrollment by age group, Dr. Toribio Bencosme Hospital, Espaillat Province, Dominican Republic, April 2021–August 2022 (n=1,255)**

**Supplementary Table 4. Univariable and multivariable odds ratios for factors associated with a positive SARS-CoV-2 virological test — Dominican Republic, 22 March 2021 – 17 August 2022**

| Variable | RT-PCR Negative, n (%) | RT-PCR Positive, n (%) | Univariate odds ratio | Multivariable odds ratio |
|---|---|---|---|---|
| Sex* | | | | |
| Female | 1095 (77.0) | 327 (23.0) | *Ref* | *Ref* |
| Male | 688 (78.4) | 189 (21.6) | 0.92 (0.75–1.13, p=0.420) | 0.93 (0.75–1.15, p=0.494) |
| Age cat, years | | | | |
| 2–17 | 376 (89.7) | 43 (10.3) | **0.37 (0.26–0.51, p<0.001)** | **0.46 (0.30–0.70, p<0.001)** |
| 18–54 | 1189 (76.4) | 368 (23.6) | *Ref* | *Ref* |
| ≥ 55 | 218 (67.3) | 106 (32.7) | **1.57 (1.21–2.03, p=0.001)** | **1.58 (1.20–2.08, p=0.001)** |
| Vaccine doses | | | | |
| None | 604 (80.1) | 150 (19.9) | *Ref* | *Ref* |
| One | 177 (74.4) | 61 (25.6) | 1.39 (0.98–1.95, p=0.060) | 1.04 (0.71–1.54, p=0.827) |
| Two | 766 (74.5) | 262 (25.5) | **1.38 (1.10–1.73, p=0.006)** | 1.09 (0.75–1.58, p=0.648) |
| Three | 230 (83.9) | 44 (16.1) | 0.77 (0.53–1.11, p=0.165) | 0.82 (0.50–1.35, p=0.441) |
| Four | 6 (100.0) | 0 (0.0) | *NC* | *NC* |
| Anti-S titer (quantile) | | | | |
| Q1 | 399 (69.4) | 176 (30.6) | *Ref* | *Ref* |
| Q2 | 431 (75.0) | 144 (25.0) | **0.76 (0.58–0.98, p=0.035)** | **0.56 (0.41–0.76, p<0.001)** |
| Q3 | 463 (80.5) | 112 (19.5) | **0.55 (0.42–0.72, p<0.001)** | **0.39 (0.27–0.56, p<0.001)** |
| Q4 | 490 (85.2) | 85 (14.8) | **0.39 (0.29–0.52, p<0.001)** | **0.27 (0.18–0.40, p<0.001)** |

Number in dataframe = 2300, Number in model = 2300, Missing = 0, AIC = 2254.3, C-statistic = 0.703, H&L = Chi-sq(8) 4.90 (p=0.768). * One sex reported as *other* not presented in table.

**Supplementary Table 5. Univariable and multivariable odds ratios for factors associated with a positive SARS-CoV-2 virological test — Pre-Delta phase of transmission, Dominican Republic, 22 March to 15 August 2021**

| Variable | RT-PCR Negative, n (%) | RT-PCR Positive, n (%) | Univariate odds ratio | Multivariable odds ratio |
|---|---|---|---|---|
| Sex | | | | |
| Female | 301 (77.8) | 86 (22.2) | *Ref* | *Ref* |
| Male | 194 (74.9) | 65 (25.1) | 1.17 (0.81–1.69, p=0.398) | 1.12 (0.75–1.66, p=0.571) |
| Age cat, years | | | | |
| 2–17 | 50 (84.7) | 9 (15.3) | 0.65 (0.29–1.29, p=0.247) | 0.59 (0.25–1.24, p=0.185) |
| 18–54 | 380 (78.2) | 106 (21.8) | *Ref* | *Ref* |
| ≥ 55 | 65 (64.4) | 36 (35.6) | **1.99 (1.24–3.13, p=0.004)** | **2.14 (1.30–3.51, p=0.003)** |
| Vaccine doses | | | | |
| None | 266 (75.8) | 85 (24.2) | *Ref* | *Ref* |
| One | 98 (76.6) | 30 (23.4) | 0.96 (0.59–1.53, p=0.860) | 1.12 (0.61–2.07, p=0.708) |
| Two | 131 (78.4) | 36 (21.6) | 0.86 (0.55–1.33, p=0.504) | 1.21 (0.58–2.46, p=0.610) |
| Three | 0 | 0 | *NC* | *NC* |
| Four | 0 | 0 | *NC* | *NC* |
| Anti–S titer (quantile) | | | | |
| Q1 | 105 (64.8) | 57 (35.2) | *Ref* | *Ref* |
| Q2 | 116 (71.6) | 46 (28.4) | 0.73 (0.46–1.17, p=0.190) | **0.45 (0.25–0.79, p=0.006)** |
| Q3 | 137 (85.1) | 24 (14.9) | **0.32 (0.19–0.55, p<0.001)** | **0.19 (0.09–0.36, p<0.001)** |
| Q4 | 137 (85.1) | 24 (14.9) | **0.32 (0.19–0.55, p<0.001)** | **0.19 (0.09–0.36, p<0.001)** |

Number in dataframe = 646, Number in model = 646, Missing = 0, AIC = 669.4, C-statistic = 0.709, H&L = Chi-sq(8) 6.51 (p=0.591).

**Supplementary Table 6. Univariable and multivariable odds ratios for factors associated with a positive SARS-CoV-2 virological test — Delta phase of transmission, Dominican Republic, 16 August 2021 to 23 December 2021**

| **Variable** | **RT-PCR Negative, n (%)** | **RT-PCR Positive, n (%)** | **Univariate odds ratio** | **Multivariable odds ratio** |
|---|---|---|---|---|
| Sex* | | | | |
| Female | 348 (73.1) | 128 (26.9) | *Ref* | *Ref* |
| Male | 205 (71.4) | 82 (28.6) | 1.09 (0.78–1.51, p=0.615) | 1.08 (0.76–1.53, p=0.673) |
| Age cat, years | | | | |
| 2–17 | 60 (78.9) | 16 (21.1) | 0.72 (0.39–1.26, p=0.264) | 0.87 (0.43–1.68, p=0.675) |
| 18–54 | 420 (72.9) | 156 (27.1) | *Ref* | *Ref* |
| ≥ 55 | 73 (65.2) | 39 (34.8) | 1.44 (0.93–2.20, p=0.098) | 1.35 (0.83–2.18, p=0.223) |
| Vaccine doses | | | | |
| None | 94 (69.6) | 41 (30.4) | *Ref* | *Ref* |
| One | 39 (61.9) | 24 (38.1) | 1.41 (0.75–2.64, p=0.282) | 2.06 (0.96–4.40, p=0.063) |
| Two | 342 (71.4) | 137 (28.6) | 0.92 (0.61–1.40, p=0.689) | 1.82 (0.92–3.59, p=0.085) |
| Three | 77 (89.5) | 9 (10.5) | **0.27 (0.12–0.56, p=0.001)** | 0.64 (0.24–1.61, p=0.353) |
| Four | 1 (100.0) | 0 (0.0) | *NC* | *NC* |
| Anti–S titer (quantile) | | | | |
| Q1 | 104 (54.5) | 87 (45.5) | *Ref* | *Ref* |
| Q2 | 142 (74.3) | 49 (25.7) | **0.41 (0.27–0.63, p<0.001)** | **0.38 (0.23–0.62, p<0.001)** |
| Q3 | 152 (79.6) | 39 (20.4) | **0.31 (0.19–0.48, p<0.001)** | **0.31 (0.19–0.51, p<0.001)** |
| Q4 | 155 (81.2) | 36 (18.8) | **0.28 (0.17–0.44, p<0.001)** | **0.32 (0.18–0.54, p<0.001)** |

Number in dataframe = 764, Number in model = 764, Missing = 0, AIC = 840.5, C-statistic = 0.714, H&L = Chi-sq(8) 6.08 (p=0.639)
* One sex reported as *other* not presented in table.

**Supplementary Table 7. Univariable and multivariable odds ratios for factors associated with a positive SARS-CoV-2 virological test — Omicron BA.1 phase of transmission, 24 December 2021 to 30 April 2022**

| Variable | RT-PCR Negative, n (%) | RT-PCR Positive, n (%) | Univariate odds ratio | Multivariable odds ratio |
|---|---|---|---|---|
| Sex | | | | |
| Female | 235 (83.3) | 47 (16.7) | *Ref* | *Ref* |
| Male | 168 (92.3) | 14 (7.7) | **0.42 (0.21-0.76, p=0.006)** | **0.42 (0.19-0.90, p=0.030)** |
| Age cat, years | | | | |
| 2–17 | 145 (98.6) | 2 (1.4) | **0.07 (0.01-0.23, p<0.001)** | 0.66 (0.06-5.17, p=0.704) |
| 18–54 | 216 (83.4) | 43 (16.6) | *Ref* | *Ref* |
| ≥ 55 | 42 (72.4) | 16 (27.6) | 1.91 (0.97-3.67, p=0.055) | 1.48 (0.63-3.47, p=0.365) |
| Vaccine doses | | | | |
| None | 135 (97.1) | 4 (2.9) | *Ref* | *Ref* |
| One | 25 (96.2) | 1 (3.8) | 1.35 (0.07-9.61, p=0.792) | 0.16 (0.01-2.31, p=0.211) |
| Two | 160 (78.8) | 43 (21.2) | **9.07 (3.56-30.71, p<0.001)** | 1.03 (0.14-8.19, p=0.975) |
| Three | 82 (86.3) | 13 (13.7) | **5.35 (1.82-19.50, p=0.004)** | 1.84 (0.25-15.16, p=0.554) |
| Four | 1 (100.0) | 0 (0.0) | *NC* | *NC* |
| Anti–S titer (quantile) | | | | |
| Q1 | 96 (82.8) | 20 (17.2) | *Ref* | *Ref* |
| Q2 | 96 (82.8) | 20 (17.2) | 1.00 (0.50-1.98, p=1.000) | 0.69 (0.27-1.76, p=0.442) |
| Q3 | 102 (87.9) | 14 (12.1) | 0.66 (0.31-1.37, p=0.268) | 0.46 (0.16-1.27, p=0.141) |
| Q4 | 109 (94.0) | 7 (6.0) | **0.31 (0.12-0.73, p=0.011)** | **0.14 (0.04-0.42, p=0.001)** |

Number in dataframe = 464, Number in model = 464, Missing = 0, AIC = 237.8, C-statistic = 0.921, H&L = Chi-sq(8) 4.05 (p=0.853)

**Supplementary Table 8. Univariable and multivariable odds ratios for factors associated with a positive SARS-CoV-2 virological test — Omicron BA.2, BA.4, BA.5 phase of transmission, 1 May 2022 to 17 August 2022**

| Variable | RT-PCR Negative, [illegible] | RT-PCR Positive, n (%) | Univariate odds ratio | Multivariable odds ratio |
|---|---|---|---|---|
| Sex | | | | |
| Female | 211 (76.2) | 66 (23.8) | *Ref* | *Ref* |
| Male | 121 (81.2) | 28 (18.8) | 0.74 (0.45-1.20, p=0.233) | 0.80 (0.45-1.38, p=0.429) |
| Age cat, years | | | | |
| 2–17 | 121 (88.3) | 16 (11.7) | **0.36 (0.19-0.64, p=0.001)** | **0.33 (0.12-0.88, p=0.030)** |
| 18–54 | 173 (73.3) | 63 (26.7) | *Ref* | *Ref* |
| ≥ 55 | 38 (71.7) | 15 (28.3) | 1.08 (0.54-2.07, p=0.812) | 1.18 (0.55-2.43, p=0.668) |
| Vaccine doses | | | | |
| None | 109 (84.5) | 20 (15.5) | *Ref* | *Ref* |
| One | 15 (71.4) | 6 (28.6) | 2.18 (0.71-6.10, p=0.150) | 1.07 (0.24-4.40, p=0.931) |
| Two | 133 (74.3) | 46 (25.7) | **1.88 (1.07-3.43, p=0.033)** | 1.09 (0.30-3.83, p=0.894) |
| Three | 71 (76.3) | 22 (23.7) | **1.69 (0.86-3.34, p=0.128)** | 0.97 (0.29-3.26, p=0.965) |
| Four | 4 (100.0) | 0 (0.0) | *NC* | *NC* |
| Anti–S titer (quantile) | | | | |
| Q1 | 73 (68.2) | 34 (31.8) | *Ref* | *Ref* |
| Q2 | 80 (74.8) | 27 (25.2) | 0.72 (0.40-1.31, p=0.290) | **0.59 (0.30-1.15, p=0.123)** |
| Q3 | 87 (82.1) | 19 (17.9) | **0.47 (0.24-0.88, p=0.021)** | **0.30 (0.14-0.62, p=0.001)** |
| Q4 | 92 (86.8) | 14 (13.2) | **0.33 (0.16-0.64, p=0.002)** | **0.24 (0.10-0.53, p=0.001)** |

Number in dataframe = 426, Number in model = 426, Missing = 0, AIC = 412.2, C-statistic = 0.764, H&L = Chi-sq(8) 8.77 (p=0.362)

**Supplementary Figure 4. SARS-CoV-2 anti-S titers by days post-symptom onset, Dominican Republic, March 2021 - May 2022.** Top plots represent anti-S titer on the y-axis and the number of days after symptom onset that the sample was collected on the x-axis, stratified by NAAT status. LOESS smoothed line and 95% CI marked with a blue line with gray shading respectively. Jittered dots represent individual study participants. No clear trend is observed. Bottom plots reflect plots above but stratified by number of COVID-19 vaccine doses received. A trend to an increase in titers is observed among NAAT positive individuals who received two or three vaccine doses, although the small number of participants makes clear interpretation challenging.

**Supplementary Table 9. Multivariable odds ratios for a positive SARS-CoV-2 virological test by anti-S quartile and number of days from symptom onset to sample collection (N = 2,300)**

| Anti-S titer, quartile | DPSO 0–4 days (n=1,541) | DPSO ≥4 days (n= 759) |
|---|---|---|
| Q1 | *Ref* | *Ref* |
| Q2 | 0.56 (0.38-0.81, p=0.002) | 0.35 (0.19-0.63, p=0.001) |
| Q3 | 0.32 (0.21-0.49, p<0.001) | 0.47 (0.23-0.93, p=0.031) |
| Q4 | 0.21 (0.13-0.33, p<0.001) | 0.37 (0.17-0.79, p=0.011) |

Ref = reference. Odds ratios with 95% CIs calculated using binomial multivariable logistic regression models with data presented for log-adjusted anti-S titers stratified by quartile. Quartiles calculated using the *quantile* function in R. Model covariates include number of COVID-19 vaccine doses received, days since last COVID-19 vaccine dose, anti-S titers, sex, age, and month of sample collection.
