## Supplementary figures and images for "Integrated SARS-CoV-2 serological and virological screening across an acute fever surveillance platform to monitor temporal changes in anti-spike antibody levels and risk of infection during sequential waves of variant transmission — Dominican Republic, March 2021 to August 2022"

### Supplementary Figure 2

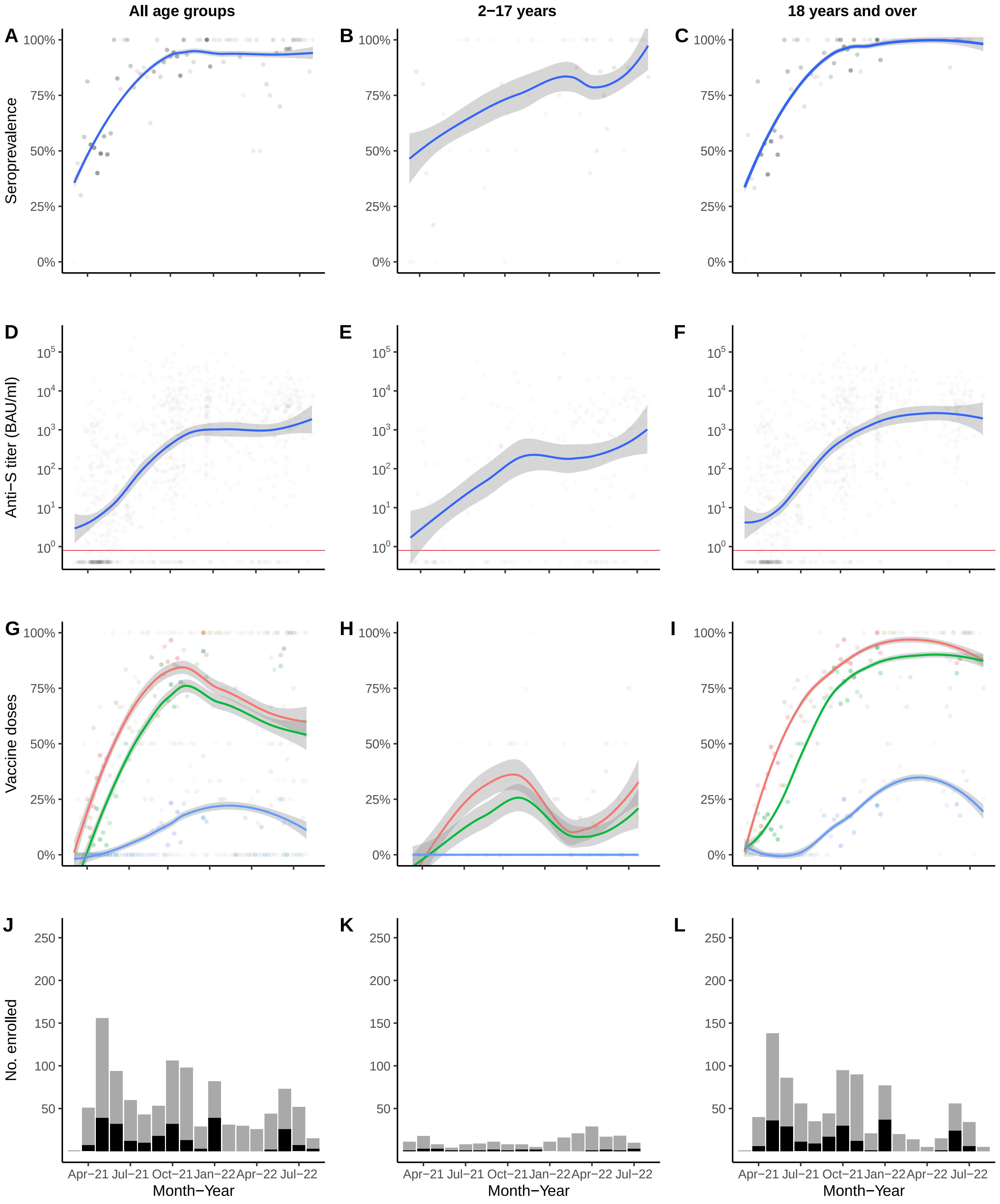

### Supplementary Figure 3

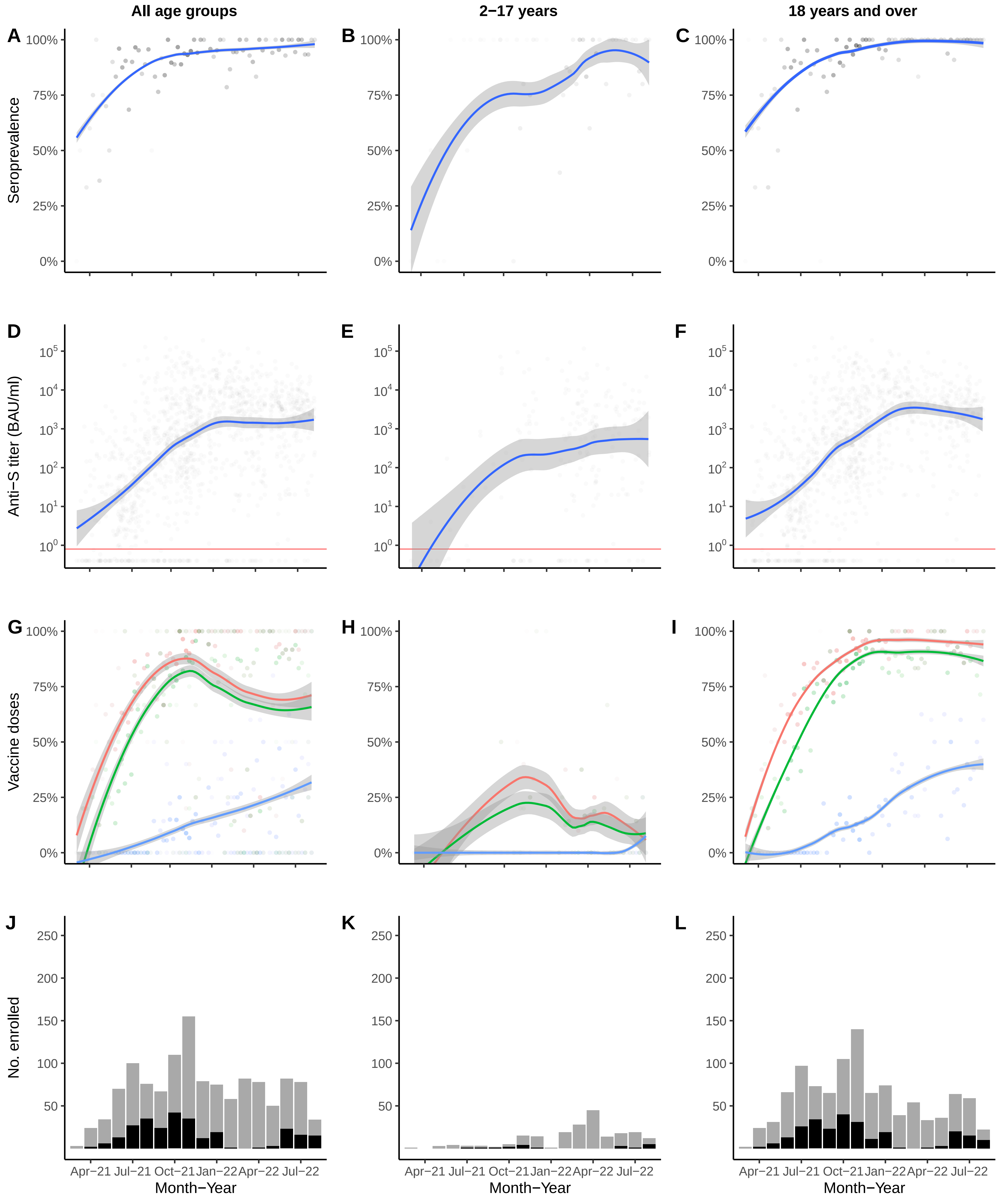

### Supplementary Figure 4

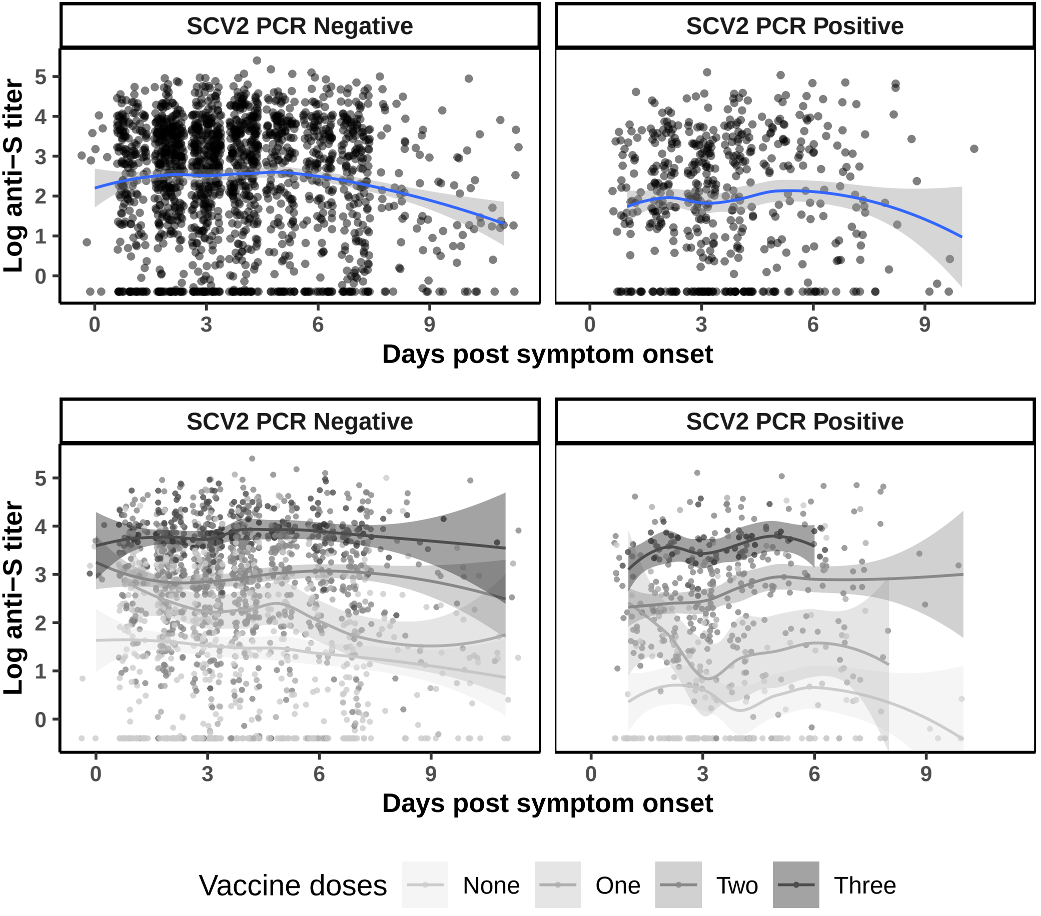
